# Explainable AI Predicts Hematoxicity from Cancer Treatment Using Multimodal Real-World Data

**DOI:** 10.64898/2026.04.29.26352032

**Authors:** Julius Keyl, Philipp Keyl, Tim Lenfers, René Hosch, Niklas Kiermeyer, Simon Schallenberg, Moon Kim, Sebastian Bauer, Nikolaos Bechrakis, Michael Forsting, Dagmar Führer-Sakel, Sied Kebir, Viktor Grünwald, Boris Hadaschik, Johannes Haubold, Ken Herrmann, Stefan Kasper, Rainer Kimmig, Stephan Lang, Tienush Rassaf, Alexander Roesch, Dirk Schadendorf, Jens T. Siveke, Martin Stuschke, Ulrich Sure, Matthias Totzeck, Anja Welt, Marcel Wiesweg, Jan Egger, Sylvia Hartmann, Grégoire Montavon, Felix Nensa, Klaus-Robert Müller, Martin Schuler, Jens Kleesiek, Frederick Klauschen

## Abstract

Adverse drug effects remain a major barrier to safe and effective cancer therapy, underscoring the need for tools that predict treatment-related toxicities. We analyzed multimodal real-world data from 14,596 cancer patients across 38 cancer entities, encompassing 330 clinical, tumor, and imaging characteristics, along with 89 anticancer agents. Hematological adverse events (HAE), defined by nadirs of hemoglobin, leukocyte, neutrophil, and platelet values within two months of treatment initiation, were highly prevalent (87.7%; 33.1% severe). We developed ***Toxix***, an explainable artificial intelligence (xAI) framework modeling interactions between patient characteristics and drug combinations. *Toxix* achieved strong predictive performance for severe toxicities (median AUROC 0.85 for anemia; >0.76 for leukopenia, neutropenia, and thrombocytopenia) and was validated in an external cohort of 2,768 patients with non-small cell lung cancer. Model explainability enabled systematic characterization of drug-patient interactions underlying HAEs. *Toxix* provides a real-world informed framework for personalized and toxicity-aware cancer therapy planning.

## Introduction

In recent years, the therapeutic landscape of oncology has expanded rapidly, with the emergence of numerous novel combinations of chemotherapy, immunotherapy, and targeted agents. These advances have led to improved survival in major cancer entities but also introduced new challenges, particularly in the management of toxicities.^1,2^ Still, cytotoxic chemotherapy is the mainstay of systemic therapy in many cancer entities, and hematologic adverse events (HAE), including anemia, neutropenia, and thrombocytopenia, are frequently observed. These HAE can require treatment interruptions or discontinuation, decreasing dose intensity and efficacy of therapy. In some cases, HAE can be life-threatening.^3–5^ There are substantial advancements in cancer therapy at the level of study populations. While the incidence of HAE is well defined at the population level, the individual prediction of toxicity would be of particular importance once a new systemic treatment is initiated.

While prediction models for treatment response are widely developed and advancing towards clinical use, tools for anticipating toxicities remain underdeveloped, leaving a critical blind spot in cancer care. Today, large volumes of multimodal real-world data (RWD) are routinely collected during cancer care. This has great potential to comprehensively characterize each patient and guide clinical decision-making. However, these data remain underutilized in clinical reality due to insufficient integration and computational leverage for treatment decision support.^6–9^ As a result, assessment of patient-specific toxicity risk continues to rely primarily on clinical experience, crude assessment of patient fitness and organ functions, and reaction to the toxicities observed following exposure of the patient.^10^ This may expose the patient to higher toxicites, which negatively impact quality of life and possible treatment efficacy. Therefore, there is a high demand for predictive methods to assess patient-specific risks and guide therapy selection and dosing.

Artificial intelligence (AI), particularly in the form of explainable AI (xAI), offers an opportunity to address this gap.^11,12^ xAI models can integrate complex patient data, uncover prognostic patterns across modalities, and deliver interpretable predictions to support clinical decision-making.^6,13–15^ Applied to pan-cancer RWD, such models have the potential to capture cross-entity patterns, arising from pathogenetic similarities and treatment protocols, providing interpretable predictions that extend beyond trial settings.^13^

Previous studies on HAE have been typically limited to single cancer types or regimens within the controlled setting of clinical trials, restricting their generalizability and failing to reflect the true incidence of HAE in routine practice.^10,16^ To address this gap, we assembled a large multimodal RWD cohort of 14,596 patients across 38 solid cancer types treated with a wide array of established and emerging cancer drug combinations. On this foundation, we developed ***Toxix***, an explainable AI framework that models patient-treatment interactions to predict HAE. By integrating 330 patient characteristics, including laboratory and imaging-derived features, *Toxix* provides both robust predictions and mechanistic insight at the cohort and individual patient level. We introduce the first adaptation of the BiLRP method, originally developed for similarity networks, to explain outcome predictions through interactions between patient features and specific therapeutic substances.^17^

This study shows that xAI applied to rich RWD can predict patient-specific toxicity and provide actionable insights to guide safer, personalized therapy.

## Results

### Dataset description

We retrospectively queried our database for cancer patients who received cancer drug therapy at the West German Cancer Center of the University Hospital Essen, one of Germany’s largest academic comprehensive cancer centers. We prespecified leukocyte, neutrophil, and platelet counts, and the hemoglobin concentration as the relevant parameters for hematologic toxicity. From a total of 150,079 cancer patients of all stages, 14,596 met the inclusion criteria, defined as availability of clinical baseline data and at least one hematologic marker (hemoglobin, neutrophils, leukocytes, platelets) measured within 60 days after treatment initiation (**Fig. 1 + Suppl Fig. 1**). Patients of this cohort were treated between April 2007 and July 2022 (median: December 2016). The most frequent cancer entities were lung cancer (n=4,139), sarcoma (n=1,501), and head and neck cancer (n=998; **Suppl. Table 1**). The collected patient data comprised 330 parameters at the time of the first drug-based cancer therapy, including laboratory values, tumor characteristics, and CT-derived body composition (Methods). In total, 89 different anticancer agents across the first recorded line of treatment had been administered in this cohort.

**Figure 1:**
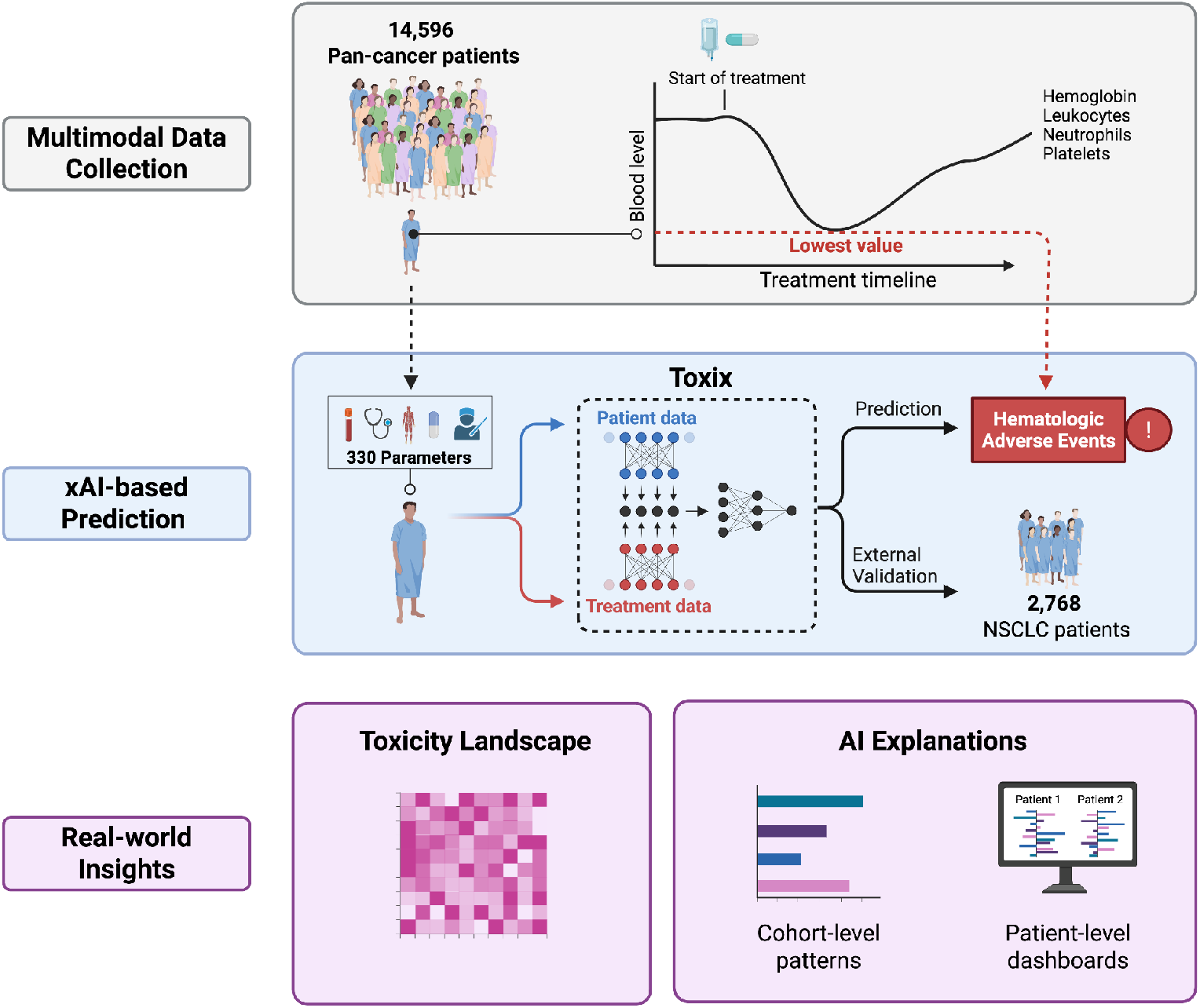
Overview of the study design. Multimodal real-world data from 14,596 patients across 38 solid tumor types were collected at the start of drug treatment. Hematologic adverse events (HAE) were defined by nadirs of hemoglobin, leukocytes, neutrophils, and platelets within 60 days of treatment initiation. These data were used to develop *Toxix*, an explainable AI framework modeling patient–treatment interactions to predict HAE and decipher the toxicity landscape at both the cohort and individual patient levels.

### Changes in hematologic parameters following treatment initiation

At baseline (before initiation of cancer treatment at our institute), anemia was the most common cytopenia, affecting 49.8% of patients, whereas thrombocytopenia (7.9%), leukopenia (3.2%), and neutropenia (2.1%) were less frequent (**Fig. 2A**). After treatment administration, all four hematologic markers showed significant deterioration. Overall, 12,806 (87.7%) patients experienced hematotoxicity, with 33.1% classified as severe (grade 3-4) per the NCI Common Terminology Criteria for Adverse Events (CTCAE) version 5.0.

**Figure 2:**
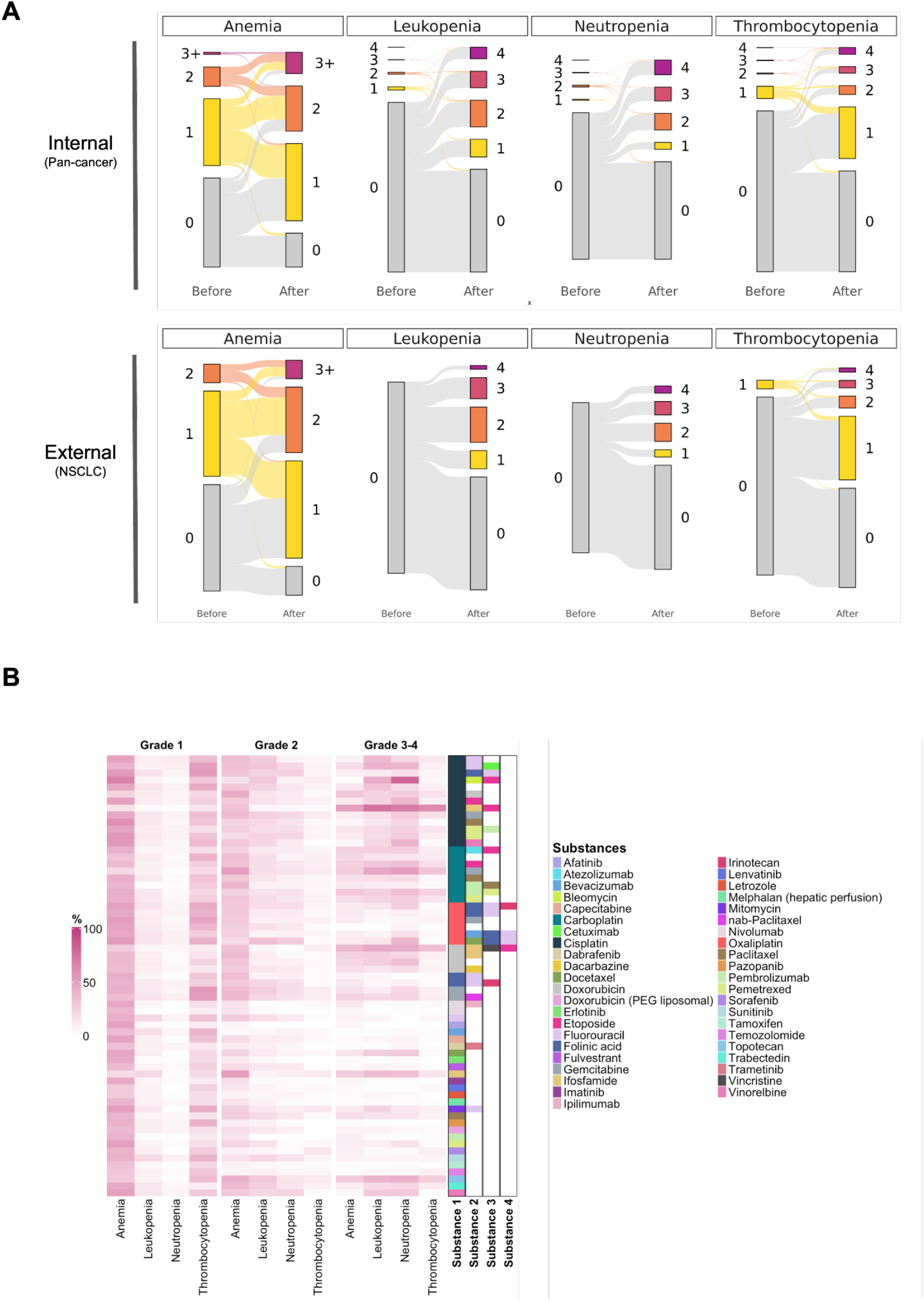
Pan-cancer landscape of HAE following cancer drug therapy. **A**: Distribution of HAE in relation to pre-treatment status. “Before” indicates the last hematologic assessment prior to therapy initiation, while ‘After’ denotes the most severe HAE recorded within two months of treatment start. **B**: Incidence of HAE within two months of cancer therapy, stratified by drug combinations. Only regimens administered to ≥50 patients are shown.

Among patients without preexisting cytopenias, new-onset events were common. Anemia developed in 64.8% (4,661/7,192) of previously non-anemic patients, with 6.3% progressing to severe grade 3-4 anemia. New-onset leukopenia occurred in 40.2% (5,510/13,695; 38.7% severe), neutropenia in 34.2% (4,046/11,836; 54.6% severe), and thrombocytopenia in 38.4% of patients (4,979/12,976; 16.6% severe).

These findings were consistent in an external cohort of patients with advanced non-small cell lung cancer (NSCLC; **Fig. 2A**). Here, 89.1% (2,465) of all patients experienced an HAE of any grade, of which 26.1% were classified as severe. As in the pan-cancer cohort, anemia was the most frequent baseline cytopenia (49.4%), followed by thrombocytopenia (4.5%). No leukopenia or neutropenia was recorded before treatment in this cohort.

### Association of hematoxicity with specific anticancer drugs and combinations

A total of 373 distinct drug combinations were administered across the entire pan-cancer cohort (**Fig. 2B**). The incidence of HAE varied markedly by regimen, with platinum-based therapies having the highest burden of hematotoxicity. The combination of cisplatin, etoposide, and ifosfamide (PEI), which is most frequently used in patients with malignant germ cell tumors, was associated with the highest rates of grade 3-4 events for anemia (50.5%), leukopenia (74.8%), and thrombocytopenia (61.7%). The combination of cisplatin, etoposide, and bleomycin (BEP), another regimen in germ cell tumor treatment, led to the highest incidence of severe neutropenia (81.5%). Interestingly, BEP resulted in relatively low rates of severe anemia (1.6%) and thrombocytopenia (4.9%).

As expected, the lowest rates of hematotoxicity were observed with orally administered targeted agents, checkpoint inhibitor monotherapy, or regionally applied therapies. For example, pazopanib was associated with the lowest incidence of severe HAE, with severe anemia and thrombocytopenia each occurring in 1.9% of patients and no cases of leukopenia or neutropenia. Similarly, low rates of severe HAE were recorded with dabrafenib plus trametinib (4.6%), regionally administered melphalan (via hepatic arteries for treatment of uveal melanoma liver metastases, 5.1%), afatinib (5.1%), and tamoxifen (6.1%).

Low rates of HAE were also observed in patients receiving pembrolizumab monotherapy (anemia: 9.2%; leukopenia: 2.1%; neutropenia: 1.6%; thrombocytopenia: 2.6%). In patients receiving pembrolizumab in combination with carboplatin and pemetrexed, as expected, HAE rates were numerically higher compared with carboplatin/pemetrexed alone, although the differences were not statistically significant (anemia: 18.9 vs 30.0%, p=0.07; leukopenia: 22.7 vs 26.2%, p=0.57; neutropenia: 25.7 vs 35.0%, p=0.17; thrombocytopenia: 14.7 vs 23.7%, p=0.11).

The marked variability in hematotoxicity across patients and treatment regimens underscored the need for predictive tools capable of anticipating individual HAE, motivating the development of an explainable AI approach to predict and interpret treatment-related toxicity presented in the following sections.

### Toxix framework for individual prediction of HAE

We developed ***Toxix***, an explainable artificial intelligence (xAI) framework that integrates rich clinical profiles described by 330 multimodal patient parameters, including tumor histology and stage, laboratory values, CT-derived body compositions, comorbidities, and procedures, to model non-linear interactions between patient characteristics and cancer drugs to predict and explain patient-specific toxicities (**Fig. 1**).^11–13,18,19^ For intravenously or regionally administered drugs, exact drug dosages were incorporated, and concurrent radiotherapy was encoded as a binary variable.

*Toxix* was trained on the full internal cohort of 14,596 pan-cancer patients to predict the occurrence of the four major HAEs (anemia, leukopenia, neutropenia, thrombocytopenia) within two months of treatment start using ten-fold cross-validation. To assess clinical utility, we report prediction performance for two endpoints: (i) any-grade complications and (ii) severe complications (CTCAE grades 3, 4). For each endpoint, we restricted evaluation to patients who had not experienced that endpoint before the start of cancer drug therapy (i.e., no complications for the any-grade analysis; no severe complications for the severe analysis).

Across all drug combinations, *Toxix* demonstrated robust predictive performance for HAE of any grade, with a median AUROC of 0.81 for anemia, 0.78 for leukopenia, 0.77 for neutropenia, and 0.73 for thrombocytopenia (**Fig. 3A**, top). For the prediction of severe HAE, Toxix showed even higher median AUROCs (Anemia: 0.85, Leukopenia: 0.79, Neutropenia: 0.76, Thrombocytopenia: 0.79). These results indicate that *Toxix* effectively captured toxicity patterns across different cytotoxic combination therapies.

**Figure 3:**
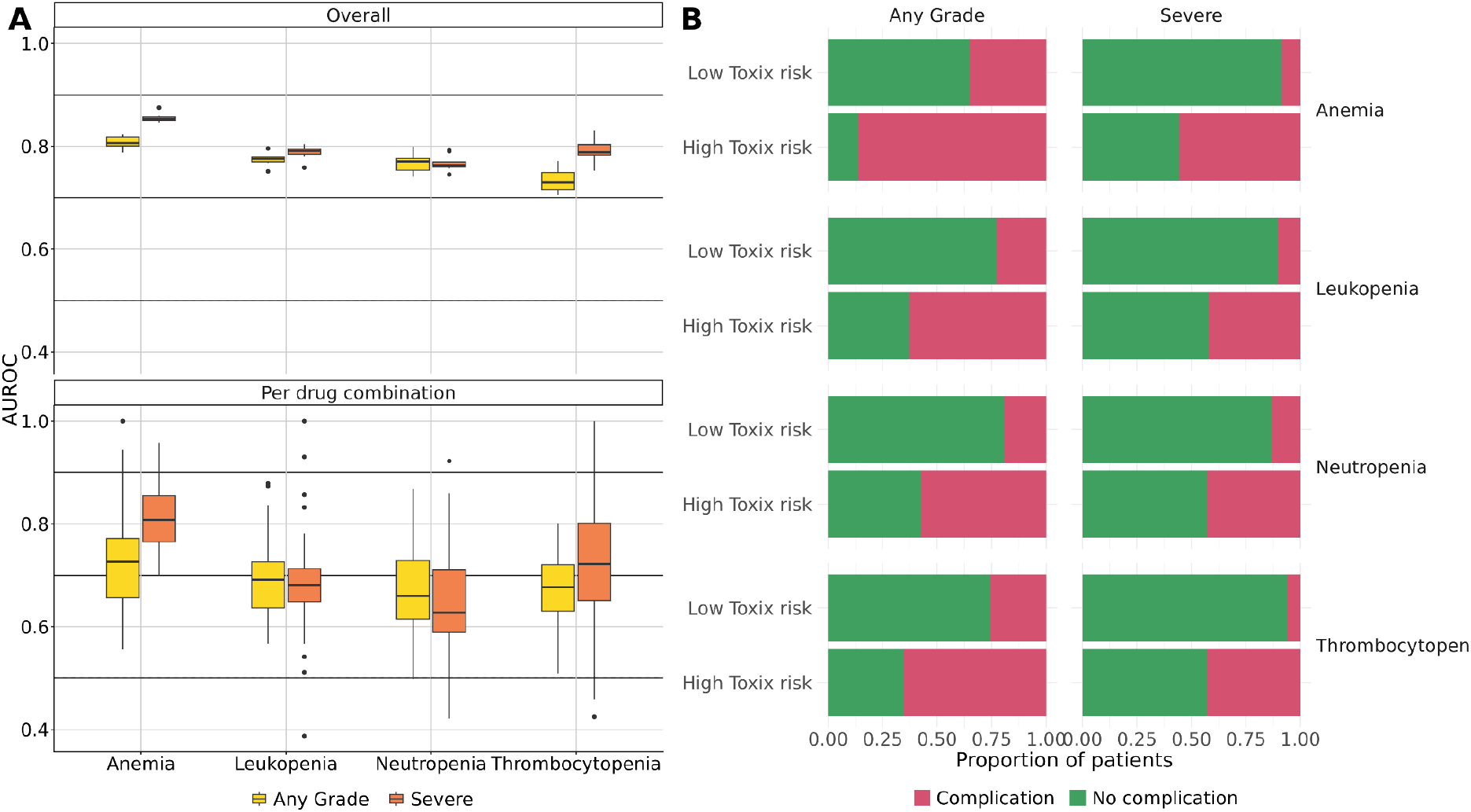
Predictive performance of *Toxix* in the internal dataset. **A:** AUROC values for prediction of HAE in the internal cohort shown overall (top) and evaluated separately for each drug combination (bottom). **B**: Positive and negative predictive values in the internal cohort. Patients were classified as high risk (top) or low risk (bottom) for developing the indicated HAE.

### Toxix performance across drug combinations

To assess clinical utility at the level of combination therapies, we evaluated *Toxix*’s performance for each drug combination individually (**Fig. 3A**, bottom). Across all combinations, *Toxix* achieved a median AUROC of 0.81 for severe anemia, 0.68 for leukopenia, 0.63 for neutropenia, and 0.72 for thrombocytopenia, although performance varied markedly between different treatment regimens (**Fig. 3A**).

Importantly, above our chosen threshold of regimens administered to at least 100 patients, predictive performance was not correlated with sample size, suggesting that differences reflected regimen-specific toxicity rather than data availability.

From a clinical perspective, patients predicted at high risk by *Toxix* showed a significantly increased relative risk of severe HAE, with 7.2 for thrombocytopenia, 6.5 for anemia, 4.1 for neutropenia, and 3.3 for leukopenia (**Fig. 3B**). For any-grade events, relative risks ranged from 2.4 to 3.0.

### Drug- and cancer-specific clinical applications

To identify clinically useful applications, we screened all cancer-drug combinations and prioritized those where Toxix yielded robust predictions for severe HAE, defined as statistically significant AUROC values after multiple-testing correction (**Table 1**).

**Table 1:**
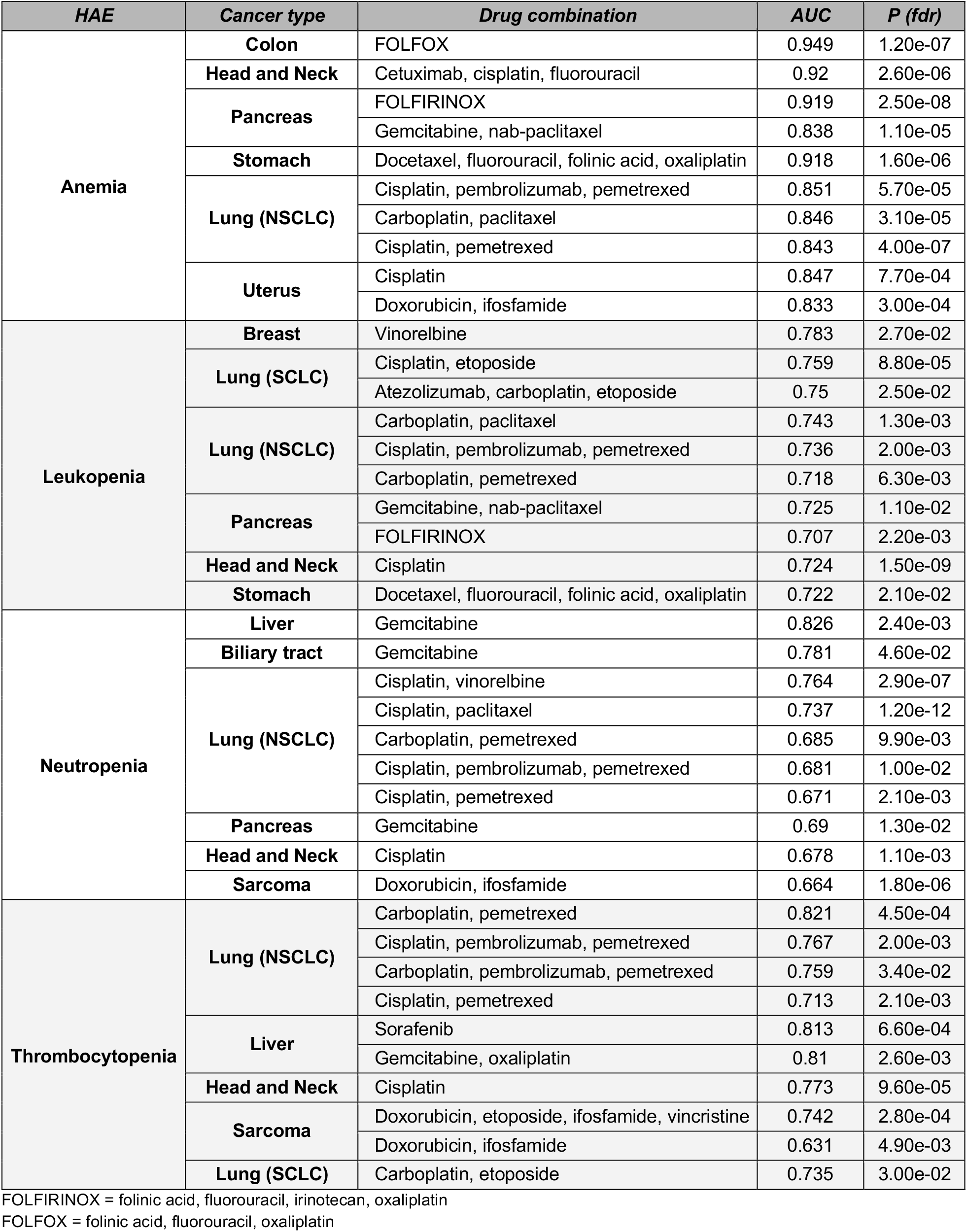
Combination therapies, stratified by cancer subtype, for which Toxix showed highest performance in predicting severe HAE. Only significant results with sufficient events are shown. Radiotherapy was included as a model feature but not used for subgroup stratification. Mann-Whitney-U test, multiple test corrected.

In Anemia, *Toxix* achieved highest AUROC for FOLFOX in colon cancer (0.95, P<0.001) and Cetuximab, cisplatin, fluorouracil in Head and neck cancers (0.92, P<0.001). For leukopenia prediction, *Toxix* showed highest predictive accuracy for Cisplatin, etoposide in SCLC (0.76, P<0.001) and for Vinorelbine in breast cancer (0.78, P=0.027). For prediction of severe neutropenia, Toxix returned the highest AUROC for Gemcitabine therapy in cancers of the liver (0.83, P=0.002) and the biliary tract (0.78, P=0.046). For thrombocytopenia prediction, highest results were achieved for Carboplatin, pemetrexed in NSCLC (0.82, P<0.001) and Sorafenib in liver cancer (0.81, P <0.001).

These entity-specific treatment scenarios represent promising candidates for future clinical validation.

### Generalization to an external validation cohort

Given *Toxix*’s robust performance in the internal cohort, we next assessed its ability to generalize to external validation data from a US nationwide electronic health record-derived database of 2,768 NSCLC patients treated with 24 different anticancer drugs. External validation is critical for clinical applicability but inherently challenging due to dataset shifts often arising from differences in patient cohorts, clinical workflows, and treatment practices.^20,21^

To adapt *Toxix* to the external validation cohort with 56 parameters, we retrained it on the internal cohort using the identical parameter set. *Toxix* predicted severe anemia with a median AUROC of 0.81, leukopenia and neutropenia with median AUROCs of 0.65 and 0.64, and thrombocytopenia with 0.78 (**Fig. 4A, B**). The prediction of HAE of any grade showed a slightly higher median AUROC for anemia (0.83), but lower for leukopenia (0.64), neutropenia (0.63) and thrombocytopenia (0.69).

**Figure 4:**
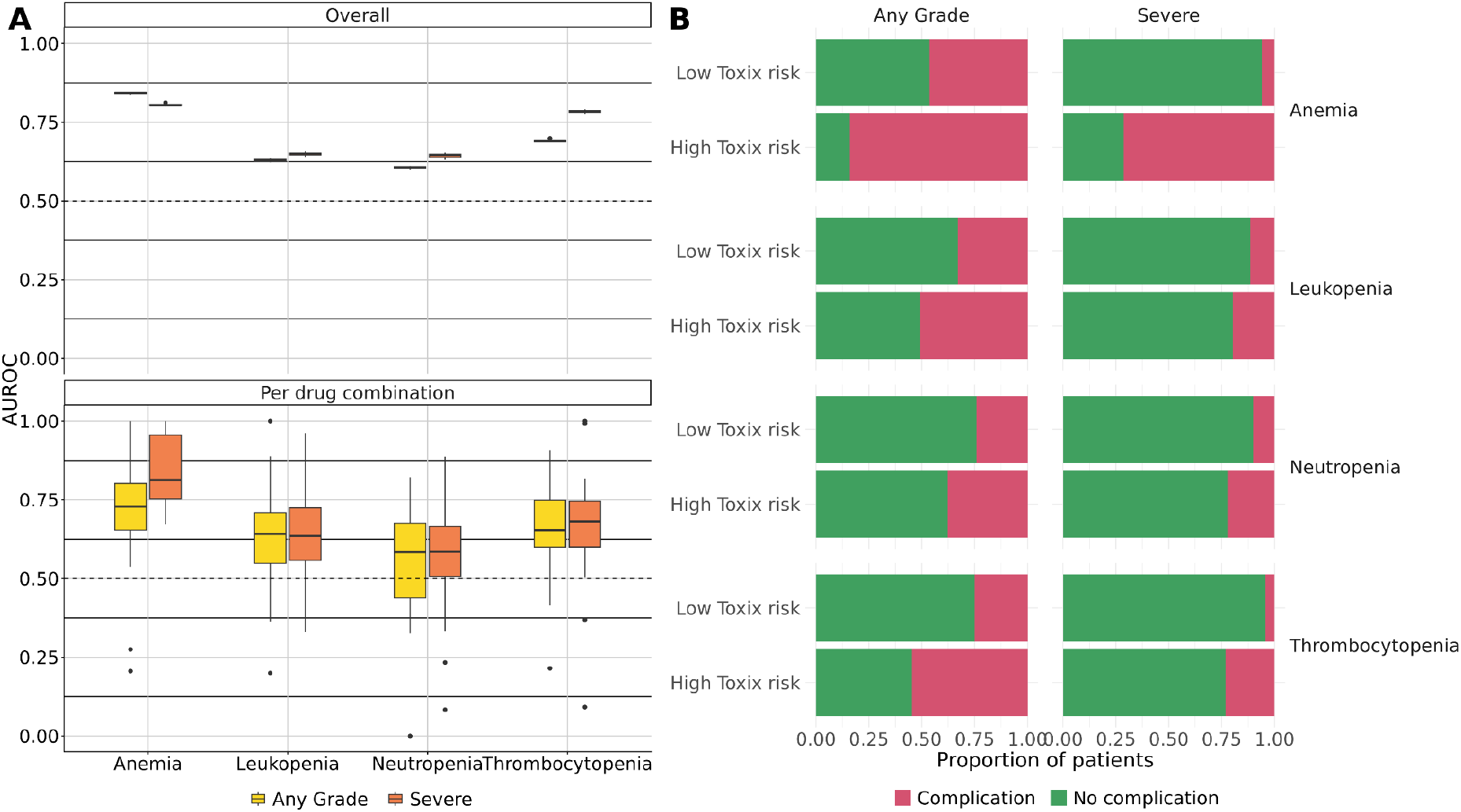
Predictive performance of *Toxix* in the external validation cohort. **A:** AUROC values for prediction of HAE in the external cohort shown overall (top) and stratified by drug combinations (bottom). **B**: Positive and negative predictive values in the external cohort. Patients were classified as high risk (top) or low risk (bottom) for developing the indicated HAE.

When stratified by individual drug combinations, *Toxix* predicted anemia with a median AUROC of 0.84 for severe anemia and 0.71 for anemia of any grade, respectively. Leukopenia was predicted with median AUROCs of 0.64 for both severe and any grade, while neutropenia showed weaker generalization (0.58 for severe and 0.57 for any grade). Thrombocytopenia showed AUROC values of 0.69 for severe HAE and 0.67 for any grade.

Importantly, patients identified by *Toxix* as high risk had a significantly elevated relative risk of severe HAE for all endpoints (anemia: 12.2, P<0.001, leukopenia: 1.7, P<0.001; neutropenia: 2.5, P<0.001; thrombocytopenia: 5.7, P<0.001).

### Identifying cohort-wide patterns contributing to HAE

Next, we investigated which patient characteristics contributed most strongly to the predicted risk of HAE. To generate patient-level explanations, we applied layer-wise relevance propagation (LRP), an xAI algorithm that attributes predictions to specific input features.^11,17,18^ For each HAE, we calculated the mean absolute LRP score (‘importance’) across patients and restricted the analysis to markers measured in at least 50% of patients to ensure stable results.

For anemia, the top-ranking markers included red blood cell distribution width and hemoglobin levels, reflecting the status of erythropoiesis, as well as prothrombin time, INR, and C-reactive protein (CRP), which indirectly influence anemia through coagulation or inflammatory pathways (**Fig. 5**). Age emerged as the dominant marker for the prediction of leukopenia, neutropenia, and thrombocytopenia.

**Figure 5:**
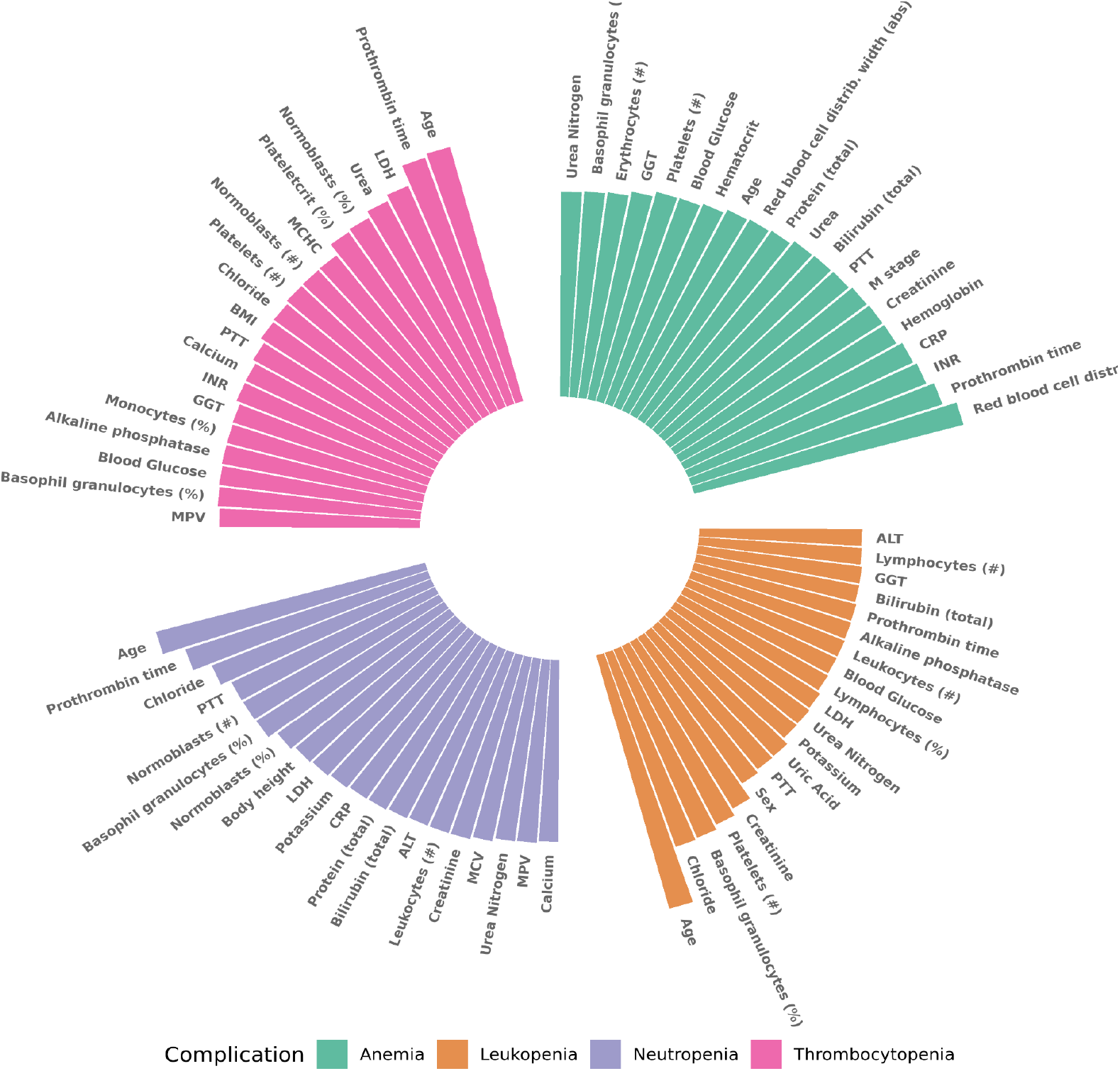
*Toxix explanations* reveal drivers of HAE. Feature importance for the prediction of anemia, leukopenia, neutropenia, and thrombocytopenia. For each HAE, the most important clinical markers are ranked from top to bottom by their contribution to model predictions. Bar length indicates the mean absolute LRP scores for each marker.

For thrombocytopenia, additional markers included prothrombin time and urea, which may be indirectly associated via coagulation or renal function. MCHC, LDH, and normoblasts likely reflect broader hematopoietic activity and platelet turnover. For neutropenia, body height was identified as an important marker. This likely reflects its role as a surrogate for body surface area, which is commonly used to calculate chemotherapy dosing and thereby links taller patients to higher absolute drug exposure. Neutropenia prediction was further associated with basophil proportion, prothrombin time, and normoblast levels.

Leukopenia prediction was most strongly linked to basophil proportion, platelet count, and creatinine, and chloride levels.

### *Toxix* disentangles patient-specific drug-toxicity interactions

Most explanation methods, including LRP, attribute model predictions to individual features without considering potential interactions between variables, in this case, patient characteristics and treatment variables. To address this limitation, we implemented an enhanced explanation technique within the Toxix framework, based on LRP and BiLRP. *Toxix* adapts the core principle of BiLRP, originally developed to identify pairs of features responsible for instance-level similarity, and extends it to multimodal interaction models to capture patient– treatment relationships (Methods). This approach enables more granular, clinically relevant, and task-specific insights.^17^ **Fig. 6A** presents a representative example of how model-derived information can be communicated to clinicians to support effective, individualized treatment guidance for cancer patients. In the first step, the “Clinician Guide” summarizes the expected changes in key markers of HAE – hemoglobin, leukocytes, neutrophils, and thrombocytes. The dashboard then attributes these predicted changes to the different cancer drugs included in the combination therapy (here, Cisplatin and Pemetrexed). Finally, the Clinician Guide provides a detailed decomposition of each predicted HAE outcome, quantifying the contribution of individual patient characteristics and administered substances to higher or lower outcome values.

**Figure 6:**
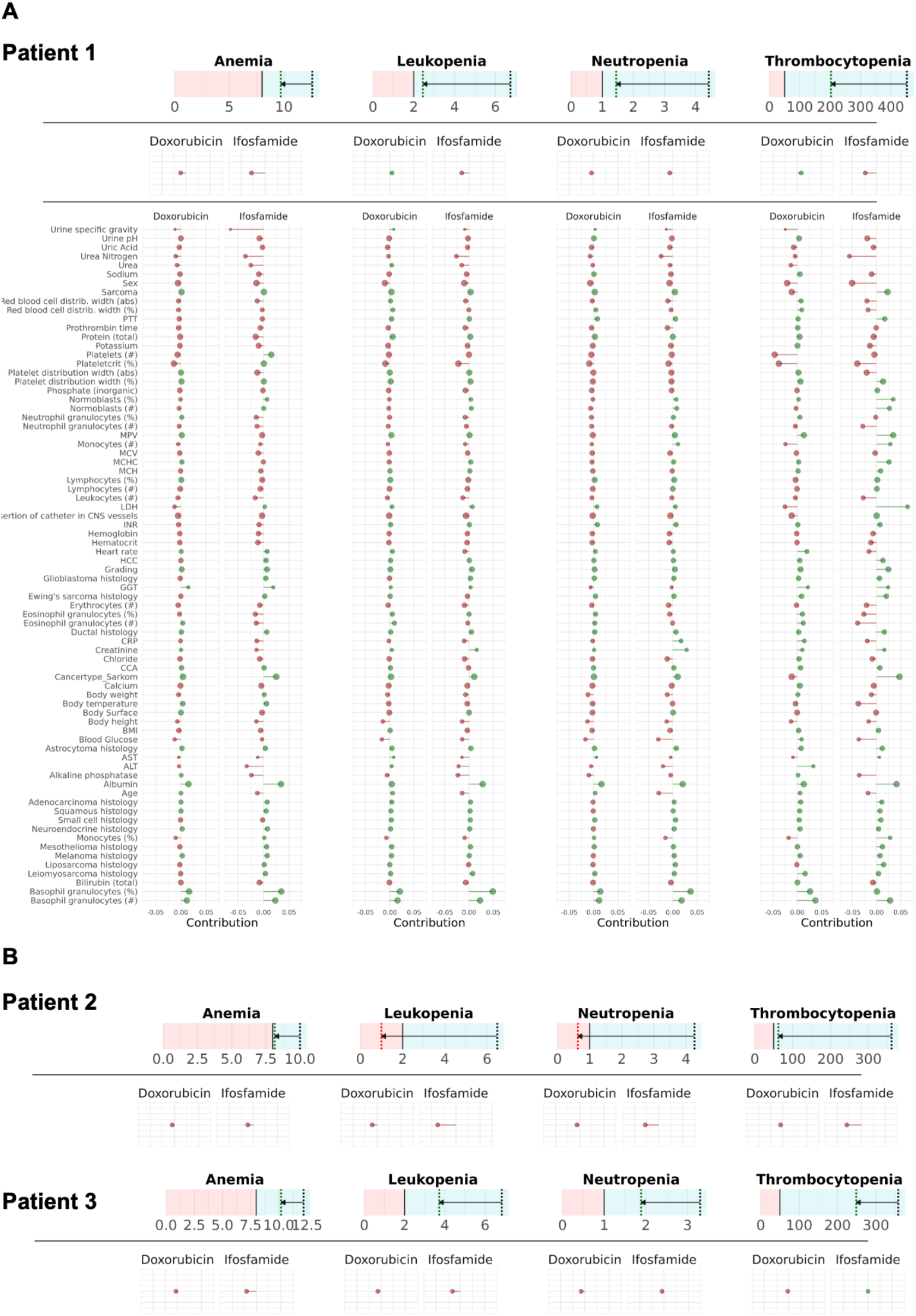
Toxix dashboard for patient-level explanations as a potential decision-support tool. **A:** For patient 1, a complete clinician guide, visualizing the predictions as well as the xAI explanations is shown. The top panel displays predicted changes from baseline values of hemoglobin, leucocytes, neutrophils, and platelets to their nadirs after treatment initiation, with arrows indicating the direction of change. The middle panel shows the estimated contributions of individual drugs to the predicted hematologic outcomes. The bottom panel further decomposes risk by highlighting interactions between specific drugs and patient characteristics. **B:** Toxicity predictions for additional example patients 2 and 3 are shown below (see Suppl. Fig. 2-3 for complete list of contributions as shown for patient 1).

**Figure 6B** shows the prediction and a high-level explanation for two additional patients. Although these patients were highly similar at baseline (all with comparable age, baseline hematologic values and treatment regimens), the model predicted markedly different outcomes. Clinicians may use this information to interpret host factors leading to HAE.

Aggregating the disentangled importances across patients by using the mean across the absolute relevance scores revealed distinct patterns of interaction between drugs and patient characteristics (**Suppl. Fig. 4, 5**). For cisplatin, oxaliplatin, doxorubicin, and paclitaxel, the prediction of thrombocytopenia was in particular driven by platelet counts, plateletcrit, and coagulation parameters such as prothrombin time and PTT. In contrast, for fluorouracil, etoposide, gemcitabine, and Ifosfamide, liver enzymes (GGT, ALT) and bilirubin emerged as highly influential predictors of thrombocytopenia. The prediction of neutropenia and leukopenia revealed highly heterogeneous importance profiles across substances. Notably, age played a dominant role for cisplatin, oxaliplatin, gemcitabine, and especially ifosfamide. For anemia prediction, different cancer drugs showed widely different important patient markers. Red blood cell distribution width was among the top five most important patient characteristics for anemia associated with cisplatin, oxaliplatin, and paclitaxel. For doxorubicin- and fluorourcil-related anemia prediction, the model was driven in particular by basophil granulocyte, eosinophil granulocyte, and neutrophil counts, respectively.

## Discussion

Hematologic adverse events (HAE) remain among the most frequent complications of cancer therapy and can significantly impact patient outcomes. In this study, we developed *Toxix*, an xAI framework trained on multimodal real-world data from 14,596 patients across 38 solid cancer types. *Toxix* predicted four key HAE, anemia, leukopenia, neutropenia, and thrombocytopenia, with high accuracy and generalized robustly to an external validation cohort of 2,768 NSCLC patients. By employing complementary explainability methods, we uncovered non-linear interactions between patient characteristics and treatment regimens, providing novel insights into the complex drivers of HAE often overlooked in the current decision-making process.

Our analysis of large-scale real-world data revealed substantial differences in HAE across drug regimens and distinct susceptibilities of individual blood cell lineages. These findings are consistent with previous studies focusing mainly on chemotherapies.^22^ By also integrating more recently emerging targeted drugs and immunotherapies, our study goes further and provides an actionable assessment of the current treatment landscape in routine clinical care. Notably, we found highly similar incidence rates of HAE between our internal dataset including multiple cancer entities, and the external validation cohort of NSCLC patients. This underscores the potential for prediction models that generalize across diverse clinical settings.

Previous attempts to predict treatment-related toxicities have typically focused on single drugs or cancer types.^23–26^ Our study demonstrates the feasibility of a pan-cancer approach, encompassing real-world patients receiving 373 different drug combinations, with each combination presenting a unique risk profile for HAE. This heterogeneity mirrors real-world practice more closely than randomized controlled trials (RCTs), where patient selection and treatment regimens are highly standardized.^27,28^ By modeling interactions between drugs and patient-specific features across cancer entities, *Toxix* reflects the complexity of real-world clinical practice, where patients often present with a wide range of comorbidities and treatment histories

Given the large number of drug combinations and the complexity of patient-specific disease contexts, we developed *Toxix* to predict the HAE risk associated with individual treatment strategies. While prediction results varied between drug combinations, *Toxix* overall showed strong predictive performance both on our internal pan-cancer dataset and an external US-wide NSCLC patient dataset, underscoring its robust generalizability across diverse clinical and geographical settings. Receiver Operating Characteristic (ROC) analysis evaluates the ranking of patients without focusing on the exact calibration of HAE, reflecting general predictive validity. However, calibration may need to be adjusted based on the clinical context. Specifically, in a real-world setting, severe HAE are relatively rare and will therefore be predicted with less likelihood. In this study, we resampled the data to balance likelihoods of hematologic adverse and non-adverse events; however, even more conservative strategies may be warranted depending on the clinical application. Although resampling strategies helped mitigate this, advances in machine learning for rare diseases and even larger multi-center cohorts will be critical to further improve robustness for these cases.^29^

Beyond prediction, our xAI approach enabled interpretable patient- and drug-level explanations, revealing both expected and novel markers of toxicity.^18^ Parameters such as age and baseline blood counts aligned with known clinical risk factors, while we found that a wide range of additional, mostly blood-based markers contribute to model predictions. These insights underscore the dual value of xAI applied to real-world data, both in enhancing clinical trust through transparency and in generating new hypotheses for translational research.^30–32^ While traditional statistical methods remain valuable for robust cohort-level analyses, they fall short in capturing these complex, non-linear interactions between a wide range of parameters and in providing patient-wise applications.

For future clinical application, the *Toxix* dashboard could serve as a practical decision-support tool to assist oncologists in managing treatment-related toxicity. When a patient is predicted to be at elevated risk for HAE, clinicians may consider adapting the therapeutic regimen—for example, by reducing the dosage, extending the interval between administrations, or temporarily deferring treatment. Moreover, the integration of explainable artificial intelligence (xAI) may facilitate the identification of transient, patient-specific factors that modulate risk, thereby enabling clinicians to postpone therapy until the patient’s condition becomes more favorable.

Despite the promising results, several limitations should be acknowledged. First, *Toxix* is designed as a decision-support tool rather than a replacement for clinical judgment. Predictions must be interpreted in the context of patient preferences and overall treatment goals.^19^ Clinical studies will be needed to evaluate the benefit of *Toxix* in real-world clinical practice in supporting treatment decisions. Second, while real-world data provide scale and diversity, they are inherently heterogeneous. Despite rigorous preprocessing and regularization strategies, residual confounding can not be completely ruled out. Real-world studies will therefore not replace RCT, but they offer potential for additional insights and hypotheses. Although RCTs in predefined patient populations remain the gold standard for establishing causality and treatment efficacy, models trained on real-world data may be better suited for deployment in clinical practice, where heterogeneity is the norm. Future studies should consider hybrid approaches that integrate both RCT and RWD sources to maximize validity and applicability

In conclusion, our study demonstrates the potential of xAI to enhance insightful patient management by modeling hematological toxicities in cancer treatment. By providing transparent personalized risk predictions, *Toxix* could help clinicians optimize treatment plans and uncover underlying complex parameter relationships. Ongoing validation and refinement of AI-based tools are essential to ensure their safe and effective implementation in diverse clinical settings.

## Methods

### Study design

The smart hospital information platform (SHIP) of University Hospital Essen was used to retrospectively evaluate 150,079 pan-cancer patients for inclusion in this study. In the final cohort, we included 14,596 adult patients who received drug-based anticancer treatment between April 2007 and July 2022 and for whom at least one measurement of hemoglobin, leukocytes, neutrophils, or platelets was available in the 60 days after the start of therapy. HAE were defined according to the Common Terminology Criteria for Adverse Events (CTCAE). The study was approved by the Ethics Committee of the Medical Faculty of the University of Duisburg-Essen (No. 21-10347-BO).

### Data acquisition

At University Hospital Essen, medical data are stored in SHIP using the FHIR format. Based on specific queries, we first retrieved all patients with solid tumors based on ICD codes (C00-C75) and identified patients who received drug-based cancer treatment and who met further criteria. The entire process is depicted in **Suppl. Fig. 1**. Subsequently, all additional data for the identified pan-cancer cohort of 14,596 patients were retrieved through SHIP. We defined specific time windows relative to the start of therapy for querying the parameter sets. This approach was chosen to find a balance between obtaining the most current and the most complete data set possible. An overview of the queried parameter sets is provided below, including the time windows where applicable:

#### *Cancer treatment* (first recorded in SHIP)

Data on first-line anticancer agents administered at the West German Cancer Center were retrieved. Agents given to fewer than 100 patients were aggregated into two categories (oral and non-oral). Oral drugs were encoded as binary variables (0: not given, 1: given), whereas dosages were used as input to the model for non-oral drugs. Similarly, radiotherapy was encoded as a binary variable. In total: 37 parameters.

#### Patient demographics

Age, sex. In total: Two parameters.

#### Body composition

Height, weight, body surface area, body mass index (BMI), and CT-derived measures of body composition. The latter were automatically extracted from abdominal CT images obtained within two months before treatment initiation and comprised muscle, bone, and fat volumes (subcutaneous, visceral, intermuscular, and total adipose tissue).^33^ To ensure comparability between patients, extracted markers were normalized by the number of abdominal CT slices. In total: Ten parameters.

#### Cancer entity

The cancer type for which patients received treatment. In total: 60 parameters.

#### *Diagnoses* (any before treatment)

All ICD-10 codes (except C00-C75) documented for at least 200 patients were collected. In total: 64 parameters.

#### *Prior medical interventions* (any before treatment)

Previous interventions were identified based on the German operation and procedure classification system (OPS). All OPS codes documented in at least 200 patients were included. In total: 44 parameters.

#### *Staging* (within one year before treatment)

Structured tumor board documentation was used to obtain T, N, and M status. In total: Three parameters.

#### *Metastasis locations* (any before treatment)

In total: Nine parameters.

#### *Clinical parameters* (within two weeks before treatment)

Oxygen saturation, body temperature, heart rate, systolic and diastolic blood pressure. In total: Five parameters.

#### *ECOG performance status* (within three months before treatment)

Extracted from structured tumor board documentation. In total: One parameter.

#### *Laboratory parameters* (within two weeks before treatment)

All laboratory parameters measured in at least 20% of patients (63 parameters), along with nine additional parameters (mainly tumor markers), which were considered relevant for subgroups. In total: 71 parameters.

#### Pathology

Detailed cancer subtype beyond ICD-10 classification, histologic tumor grade, immunohistochemical results, and somatic tumor mutations. In total: 22 parameters

#### Smoking status

Smoking status and cumulative exposure (pack-years). In total: Two parameters

#### *HAE* (within 60 days after treatment initiation)

Highest recorded grade of anemia, leukopenia, neutropenia, and thrombocytopenia, as defined by the NCI Common Terminology Criteria for Adverse Events (CTCAE) version 5.0. In total: Four parameters.

### Data preprocessing

For continuous variables, outliers exceeding three standard deviations from the mean were excluded. These variables were then normalized to have a mean of zero and unit variance. Categorical variables were represented on an ordinal scale. Information on diagnoses, cancer entities, medical interventions, and oral cancer treatments was represented using binary encodings. Non-oral cancer treatment dosages were encoded as numerical variables. For further analyses and descriptive purposes, individual cancer entities were aggregated into broader categories (**Suppl. Table 1**). Missing data were handled using a feature expansion strategy to preserve interpretability: x → (x, 1-x). Missing values were encoded as (0,0) as previously described.^13–15,34^ Feature expansion was restricted to features with incomplete data.

### External Flatiron Health dataset

We used the US-based, electronic health record-derived de-identified Flatiron Health Research Database as external validation data.^35^ This study included data from 2,768 patients diagnosed with advanced non-small cell lung cancer (NSCLC) between January 2011 and October 2022. Most patients (84.4%) originated from community oncology settings. The data are de-identified and subject to obligations to prevent re-identification and protect patient confidentiality. Due to patient de-identification requirements, patients with a birth year of 1937 or earlier may have an adjusted birth year in the Flatiron datasets.

For analysis in this study, extreme outliers were first manually removed, followed by the exclusion of outliers exceeding three standard deviations from the mean. The remaining preprocessing of the data was conducted similarly to the internal dataset. The final validation dataset contained a total of 56 parameters.

### Model architecture

To predict events, we used a neural network architecture that received two inputs: a vector containing the patient characteristics and a vector providing information about the given drug combination. This vector had the dimension of the number of individual compounds. Each entry of the vector was set to the dosage of the given compound and 0 if the drug was not given.

Patient and drug combination vectors were passed through an individual linear layer, respectively, mapping into a latent space of dimension 1000. Here, the interaction was calculated using the element-wise product of the two latent vectors. Unlike additive layers, the product layer enables model responses that are based on the conjunction of patterns from the two data modalities, that is, better accounting for patient-drug interaction effects. The resulting latent vector was mapped through an MLP with one hidden layer of dimension 1000 that mapped the latent vector to the one-dimensional output.

### Model Training

We used a ten-fold cross-validation to assess the quality of the trained folds. In each fold, 10% of data were held out as test data used only to evaluate the model performance. From the remaining 90% of samples, 10% of data samples were randomly selected as validation set to stop model training.^36^ All remaining samples were used as training dataset.

Models were trained for up to 800 epochs with a learning rate of 0.01 using Stochastic gradient descent with 0.9 momentum and a batch size of 1000. The Huber loss was used instead of Mean squared error, as this robustly mitigated the occurrence of high gradients due to the product structure in this particular network architecture.^37^ We performed early stopping based on a hold out validation set using the following procedure: If the performance on the validation set did not improve for 50 epochs for the first time, the learning rate was set to 0.001. At the second time, training was stopped and the best performing model was used for testing performance on the held out test set.

We used a dropout rate of 0.5 in intermediate layers.

Diagnostic input features and labels were normalized by first fitting the sklearn Powertransformer on the training set. Drug dosages were scaled between 0 and 1 for each substance. Using the fitted parameters the train, validation and test data set were normalized. We trained a separate model for each HAE and grade to realize a consistent resampling regime for each of each configurations:

Models were trained to regress the principal marker of each HAE grade, using a resampling strategy to equally balance samples with and without HAE post-treatment. This was important in particular for rare (mostly high grade) HAE to improve sensitivity.

### Explaining ML Predictions

To explain the model’s predictions, we extended the LRP and BiLRP methods to produce explanations that uncover interactions.^17,18^ Similar to LRP, our method starts with the prediction (the value obtained at the output of the neural network), redistributes it backwards, layer after layer, by means of propagation rules until the product layer is reached. Once the product layer is reached, relevance scores are reinterpreted from contribution of single neurons to contributions of pairs of neurons (the two inputs of the product), and these pairwise contributions are propagated, similarly to BiLRP, from layer to layer until the input is reached. Computationally, we follow the strategy outlined in the BiLRP paper, where the explanation can be computed as the matrix of partial explanations.^17^ Denoting by x and x’ the two input modalities, by a and a’ the two activation vectors, by z the output of the product, and by y the output of the model, the overall explanation can be expressed as the matrix product:

E(y|x,x’) = \sum_{a,a’} y P(y|z(a,a’)) P(a|x) P(a’|x’) where the terms on the right hand side of the equation encode the proportion (“P”) of relevance that flows through the different units of the network, as determined by LRP. This equation leads to a matrix of size dim(x) x dim(x’) containing the contribution of each pair of features from the patient and drug modalities to the prediction at the output of the network, and can be interpreted as patient-drug interaction effects.

### Statistics

Statistical analyses were performed in R statistical packages.^38^ The statistical tests were two-sided and results were regarded as significant if P<0.05. Wilcoxon and Mann-Whitney-U tests were performed in the package Hmisc.^39^ Multiple testing correction was applied using the false discovery rate (FDR) method.

## Figures

Fig. 1 was created with BioRender.com and Fig. 2B with ComplexHeatmap.^40^ All other plots were created with ggplot2.^41^

## Acknowledgements

The data for this project was provided by the Smart Hospital Information Platform (SHIP), managed by the Data Integration Center at the University Medicine Essen. SHIP serves as a comprehensive digital health platform for integrating data from all major clinical subsystems using a holistic FHIR-based approach. It enables the purification, analysis, distribution, and visualization of clinical data. Computations were carried out by KI Translation Essen (KITE).

## Contributions

Conceptualization: J.Keyl and P.K.

Methodology: J.Keyl, P.K., G.M.

Formal analysis: J.Keyl and P.K.

Investigation: all authors.

Resources: J.Kleesiek, M.Schuler., F.K., K.-R.M., S.S., M.K., S.B., N.B., M.F., D.F.-S., S.Kebir, V.G., B.H., J.H., K.H., S.Kasper, R.K., S.L., T.R., A.R., D.S., J.T.S., M.Stuschke, U.S., M.T., A.W., M.W., J.E., S.H., F.N.

Data acquisition: J.Keyl, T.L., R.H.

Writing—original draft: J.Keyl and P.K.

Writing—review and editing: all authors.

Supervision: F.K., J.Kleesiek, M.Schuler and K.-R.M.

## Data Availability Statement

Anonymized data are available upon reasonable request. Data cannot be shared with investigators outside the institution without consent. The data that support the findings of this study have been originated by and are the property of Flatiron Health, Inc. Requests for data sharing by license or by permission for the specific purpose of replicating results in this manuscript can be submitted to.

## References

1. Tsimberidou AM, Fountzilas E, Nikanjam M, Kurzrock R. Review of precision cancer medicine: Evolution of the treatment paradigm. Cancer Treat Rev. 2020 Jun;86:102019.

2. Siegel RL, Giaquinto AN, Jemal A. Cancer statistics, 2024. CA: a cancer journal for clinicians. 2024;74(1).

3. Lalami Y, Klastersky J. Impact of chemotherapy-induced neutropenia (CIN) and febrile neutropenia (FN) on cancer treatment outcomes: An overview about well-established and recently emerging clinical data. Crit Rev Oncol Hematol. 2017 Dec;120:163–79.

4. Michot JM, Lazarovici J, Tieu A, Champiat S, Voisin AL, Ebbo M, et al. Haematological immune-related adverse events with immune checkpoint inhibitors, how to manage? Eur J Cancer. 2019 Nov;122:72–90.

5. Kroschinsky F, Stölzel F, von Bonin S, Beutel G, Kochanek M, Kiehl M, et al. New drugs, new toxicities: severe side effects of modern targeted and immunotherapy of cancer and their management. Critical Care. 2017 Apr 14;21(1):89.

6. Lipkova J, Chen RJ, Chen B, Lu MY, Barbieri M, Shao D, et al. Artificial intelligence for multimodal data integration in oncology. Cancer Cell. 2022 Oct 10;40(10):1095–110.

7. Keyl J, Kasper S, Wiesweg M, Götze J, Schönrock M, Sinn M, et al. Multimodal survival prediction in advanced pancreatic cancer using machine learning. ESMO Open. 2022 Oct;7(5):100555.

8. Keyl J, Hosch R, Berger A, Ester O, Greiner T, Bogner S, et al. Deep learning-based assessment of body composition and liver tumour burden for survival modelling in advanced colorectal cancer. Journal of cachexia, sarcopenia and muscle. 2023;14(1):545–52.

9. Keyl J, Bucher A, Jungmann F, Hosch R, Ziller A, Armbruster R, et al. Prognostic value of deep learning-derived body composition in advanced pancreatic cancer-a retrospective multicenter study. ESMO Open. 2024 Jan;9(1):102219.

10. Osterman CK, Sanoff HK, Wood WA, Fasold M, Lafata JE. Predictive Modeling for Adverse Events and Risk Stratification Programs for People Receiving Cancer Treatment. JCO Oncol Pract. 2022 Feb;18(2):127–36.

11. Samek W, Montavon G, Lapuschkin S, Anders CJ, Müller KR. Explaining deep neural networks and beyond: A review of methods and applications. Proceedings of the IEEE. 2021;109(3):247–78.

12. Klauschen F, Dippel J, Keyl P, Jurmeister P, Bockmayr M, Mock A, et al. Toward Explainable Artificial Intelligence for Precision Pathology. Annual Review of Pathology: Mechanisms of Disease. 2024 Jan 24;19(Volume 19, 2024):541–70.

13. Keyl J, Keyl P, Montavon G, Hosch R, Brehmer A, Mochmann L, et al. Decoding pan-cancer treatment outcomes using multimodal real-world data and explainable artificial intelligence. Nat Cancer. 2025 Feb;6(2):307–22.

14. Keyl P, Bockmayr M, Heim D, Dernbach G, Montavon G, Müller KR, et al. Patient-level proteomic network prediction by explainable artificial intelligence. NPJ Precis Oncol. 2022 Jun 7;6(1):35.

15. Keyl P, Bischoff P, Dernbach G, Bockmayr M, Fritz R, Horst D, et al. Single-cell gene regulatory network prediction by explainable AI. Nucleic Acids Res. 2023 Feb 28;51(4):e20.

16. Pasello G, Pavan A, Attili I, Bortolami A, Bonanno L, Menis J, et al. Real world data in the era of Immune Checkpoint Inhibitors (ICIs): Increasing evidence and future applications in lung cancer. Cancer Treat Rev. 2020 Jul;87:102031.

17. Eberle O, Büttner J, Kräutli F, Müller KR, Valleriani M, Montavon G. Building and interpreting deep similarity models. IEEE Transactions on Pattern Analysis and Machine Intelligence. 2022;44(3):1149–61.

18. Bach S, Binder A, Montavon G, Klauschen F, Müller KR, Samek W. On Pixel-Wise Explanations for Non-Linear Classifier Decisions by Layer-Wise Relevance Propagation. PLOS ONE. 2015 Oct 7;10(7):e0130140.

19. Stenzinger A, Alber M, Allgäuer M, Jurmeister P, Bockmayr M, Budczies J, et al. Artificial intelligence and pathology: From principles to practice and future applications in histomorphology and molecular profiling. Seminars in Cancer Biology. 2022 Sep 1;84:129–43.

20. Sugiyama M, Krauledat M, Müller KR. Covariate Shift Adaptation by Importance Weighted Cross Validation. Journal of Machine Learning Research. 2007;8(35):985–1005.

21. Sugiyama M, Kawanabe M. Machine Learning in Non-Stationary Environments: Introduction to Covariate Shift Adaptation [Internet]. The MIT Press; 2012 [cited 2025 Sep 2]. Available from: https://direct.mit.edu/books/monograph/3774/Machine-Learning-in-Non-Stationary

22. Wu Y, Aravind S, Ranganathan G, Martin A, Nalysnyk L. Anemia and thrombocytopenia in patients undergoing chemotherapy for solid tumors: a descriptive study of a large outpatient oncology practice database, 2000-2007. Clin Ther. 2009;31 Pt 2:2416–32.

23. Matsubara J, Ono M, Negishi A, Ueno H, Okusaka T, Furuse J, et al. Identification of a predictive biomarker for hematologic toxicities of gemcitabine. J Clin Oncol. 2009 May 1;27(13):2261–8.

24. Cuplov V, André N. Machine learning approach to forecast chemotherapy-induced haematological toxicities in patients with rhabdomyosarcoma. Cancers. 2020;12(7):1944.

25. Topf V, Kheifetz Y, Daum S, Ballhausen A, Schwarzer A, Trung KV, et al. Individual hematotoxicity prediction of further chemotherapy cycles by dynamic mathematical models in patients with gastrointestinal tumors. J Cancer Res Clin Oncol. 2023 Aug;149(10):6989–98.

26. Oyaga-Iriarte E, Insausti A, Sayar O, Aldaz A. Prediction of irinotecan toxicity in metastatic colorectal cancer patients based on machine learning models with pharmacokinetic parameters. J Pharmacol Sci. 2019 May;140(1):20–5.

27. Averitt AJ, Weng C, Ryan P, Perotte A. Translating evidence into practice: eligibility criteria fail to eliminate clinically significant differences between real-world and study populations. npj Digit Med. 2020 May 11;3(1):1–10.

28. Zauderer MG. Practical Application of Real-World Evidence in Developing Cancer Therapies. JCO Clin Cancer Inform. 2019 Dec;(3):1–2.

29. Dippel J, Prenißl N, Hense J, Liznerski P, Winterhoff T, Schallenberg S, et al. AI-Based Anomaly Detection for Clinical-Grade Histopathological Diagnostics. NEJM AI. 2024 Oct 24;1(11):AIoa2400468.

30. Asan O, Bayrak AE, Choudhury A. Artificial intelligence and human trust in healthcare: focus on clinicians. Journal of medical Internet research. 2020;22(6):e15154.

31. Markus AF, Kors JA, Rijnbeek PR. The role of explainability in creating trustworthy artificial intelligence for health care: A comprehensive survey of the terminology, design choices, and evaluation strategies. J Biomed Inform. 2021 Jan;113:103655.

32. Booth CM, Karim S, Mackillop WJ. Real-world data: towards achieving the achievable in cancer care. Nat Rev Clin Oncol. 2019 May;16(5):312–25.

33. Koitka S, Kroll L, Malamutmann E, Oezcelik A, Nensa F. Fully automated body composition analysis in routine CT imaging using 3D semantic segmentation convolutional neural networks. Eur Radiol. 2021 Apr 1;31(4):1795–804.

34. Lenz OU, Peralta D, Cornelis C. Polar Encoding: A Simple Baseline Approach for Classification with Missing Values. IEEE Transactions on Fuzzy Systems. 2024;

35. Flatiron Health. Database Characterization Guide. Flatiron.com. Published March 18, 2025. Accessed November 21, 2025. https://flatiron.com/database-characterization.

36. Prechelt L. Early Stopping - But When? In: Orr, GB, Müller, KR (eds) Neural Networks: Tricks of the Trade Lecture Notes in Computer Science, vol 1524 Springer, Berlin, Heidelberg. 2002.

37. Hastie T, Friedman J, Tibshirani R. The elements of statistical learning: data mining, inference, and prediction. Springer; 2009.

38. R Core Team. R: A language and environment for statistical computing. R Foundation for Statistical Computing. 2022;

39. Harrell Jr FE, Dupont C. Package ‘hmisc’. CRAN2018. 2019;2019:235–6.

40. Gu Z, Eils R, Schlesner M. Complex heatmaps reveal patterns and correlations in multidimensional genomic data. Bioinformatics. 2016 Sep 15;32(18):2847–9.

41. Wickham H. Data analysis. ggplot2: elegant graphics for data analysis. 2016;189–201.

