## Supplementary Material for "Explainable AI Predicts Hematoxicity from Cancer Treatment Using Multimodal Real-World Data"

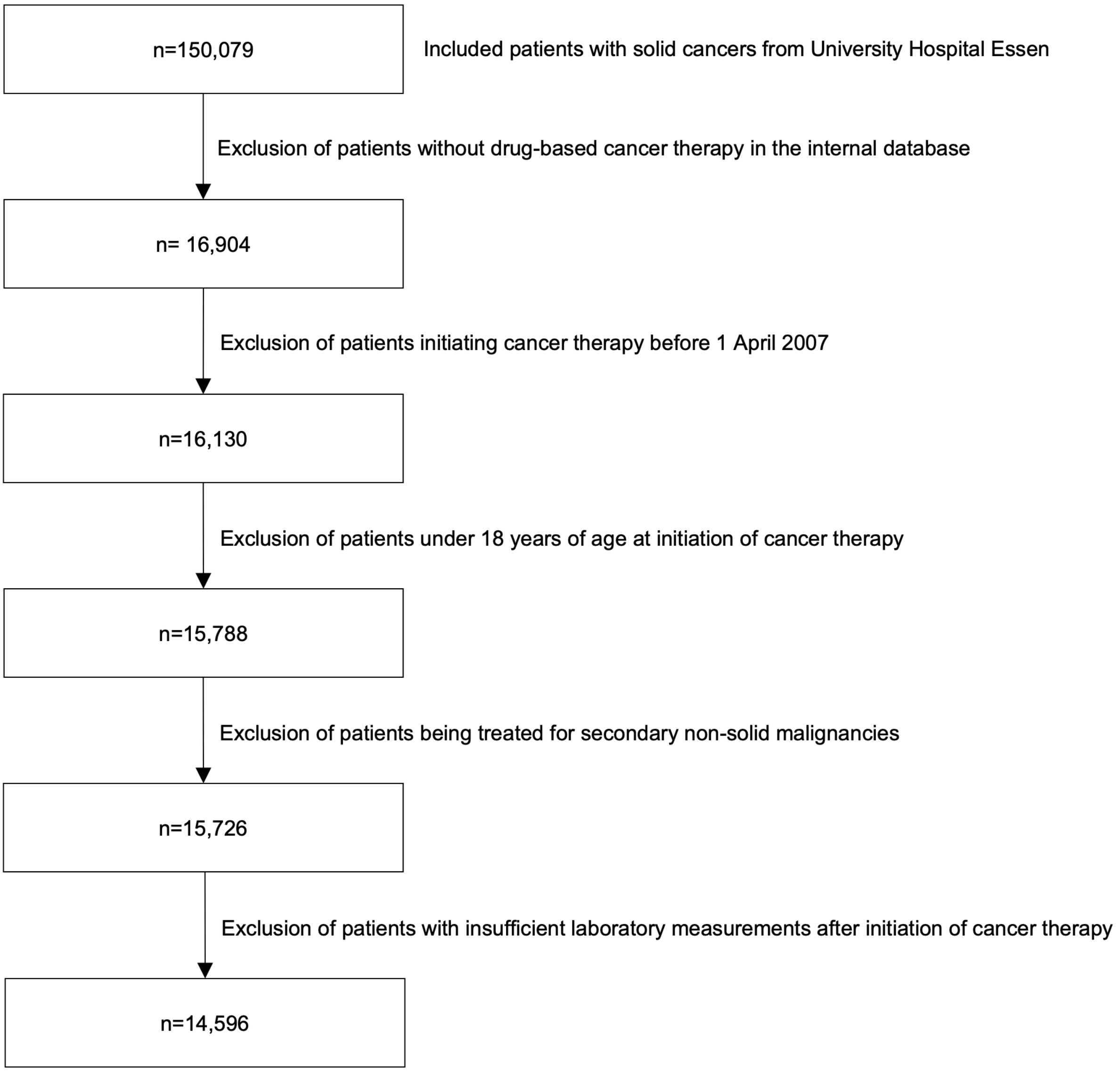


**Supplementary Figure 1: Flowchart depicting the patient enrollment process**

**Supplementary Table 1: Description of the patient cohort**

| Cancer | ICD | Patients |
|---|---|---|
| Lung | C34 | 4,139 |
| Sarcoma | C40, C41, C47-C49 | 1,501 |
| Head and Neck | C01-C13, C30-C32 | 998 |
| Breast | C50 | 974 |
| Liver | C22 | 684 |
| Colon | C18 | 585 |
| Melanoma | C43 | 580 |
| Pancreas | C25 | 579 |
| Brain | C71 | 520 |
| Stomach | C16 | 517 |
| Esophagus | C15 | 403 |
| Rectum | C20 | 359 |
| Eye | C69 | 350 |
| Kidney | C64, C65 | 277 |
| Uterus | C53-C55 | 259 |
| Testis | C62 | 239 |
| Mesothelioma | C45 | 217 |
| Skin | C44 | 207 |
| Prostate | C61 | 171 |
| Biliary tract | C24 | 156 |
| Bladder | C67 | 147 |
| Ovary | C56 | 125 |
| Thyroid gland | C73 | 96 |
| Heart | C38 | 81 |
| Rectosigmoid junction | C19 | 76 |
| Small intestine | C17 | 70 |
| Gallbladder | C23 | 63 |
| Anus | C21 | 45 |
| Urethra | C68 | 32 |
| Other endocrine gland | C75 | 31 |
| Vulva | C51 | 29 |
| Other digestive organs | C26 | 20 |
| Thymus | C37 | 20 |
| Adrenal gland | C74 | 13 |
| Penis | C60 | 11 |
| Ureter | C66 | 10 |
| Female genital | C57 | 8 |
| Central nervous system | C72 | 4 |

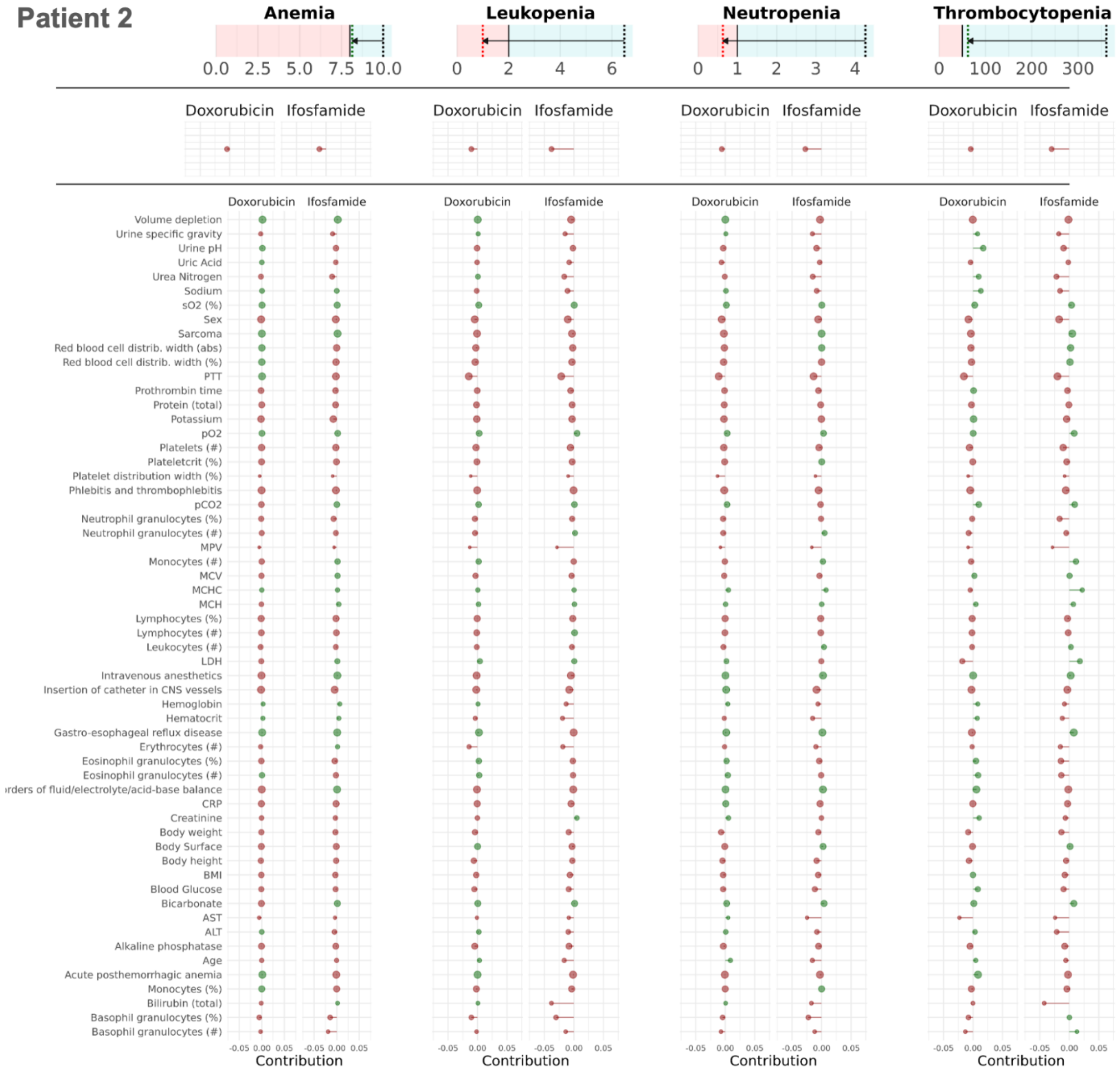


**Supplementary Figure 2:** Clinican Guide for Patient 2. The top panel displays predicted changes from baseline values of hemoglobin, leucocytes, neutrophils, and platelets to their nadirs after treatment initiation, with arrows indicating the direction of change. The middle panel shows the estimated contributions of individual drugs to the predicted hematologic outcomes. The bottom panel further decomposes risk by highlighting interactions between specific drugs and patient characteristics.

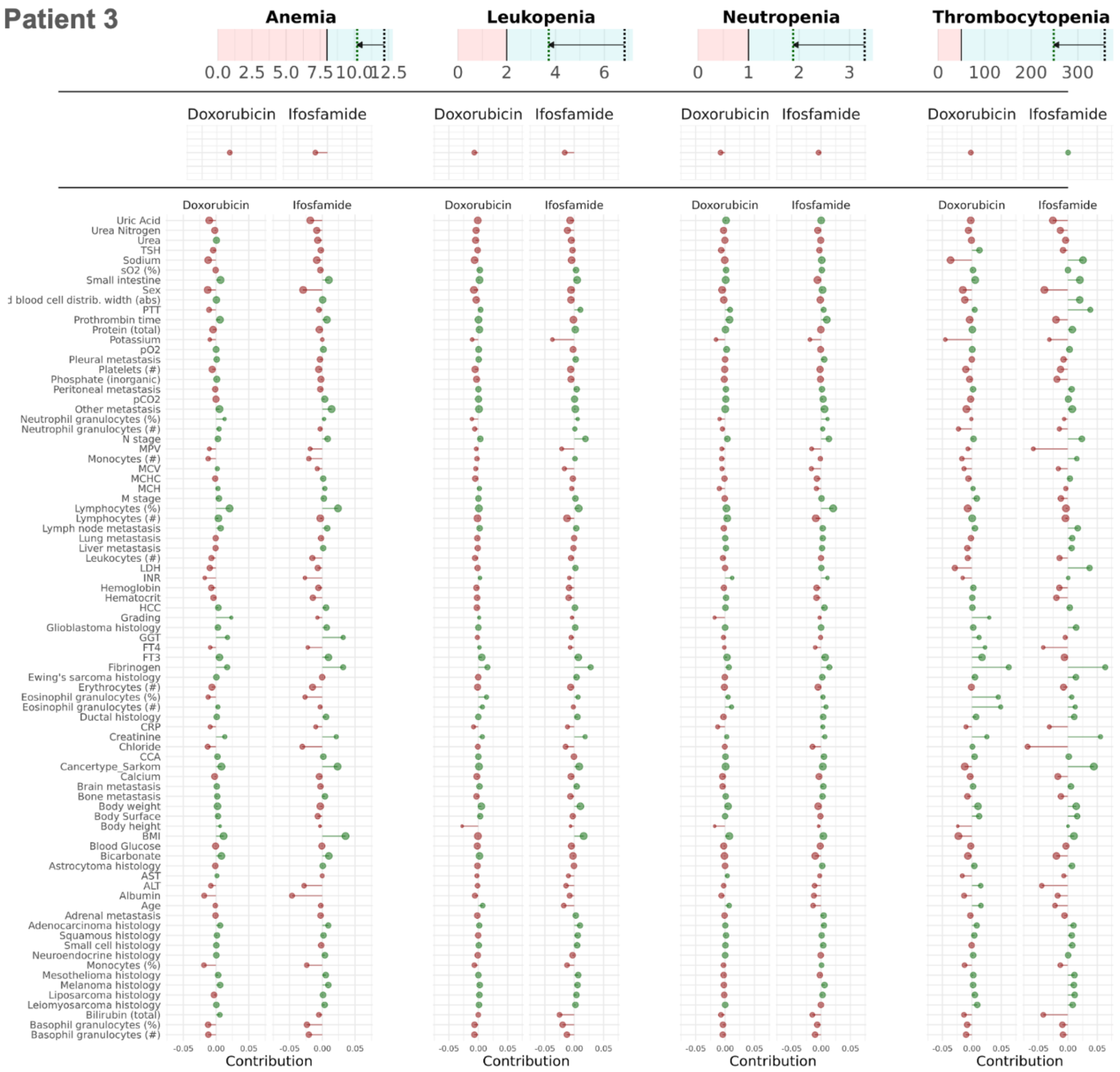


**Supplementary Figure 3:** Clinican Guide for Patient 3. The top panel displays predicted changes from baseline values of hemoglobin, leucocytes, neutrophils, and platelets to their nadirs after treatment initiation, with arrows indicating the direction of change. The middle panel shows the estimated contributions of individual drugs to the predicted hematologic outcomes. The bottom panel further decomposes risk by highlighting interactions between specific drugs and patient characteristics.

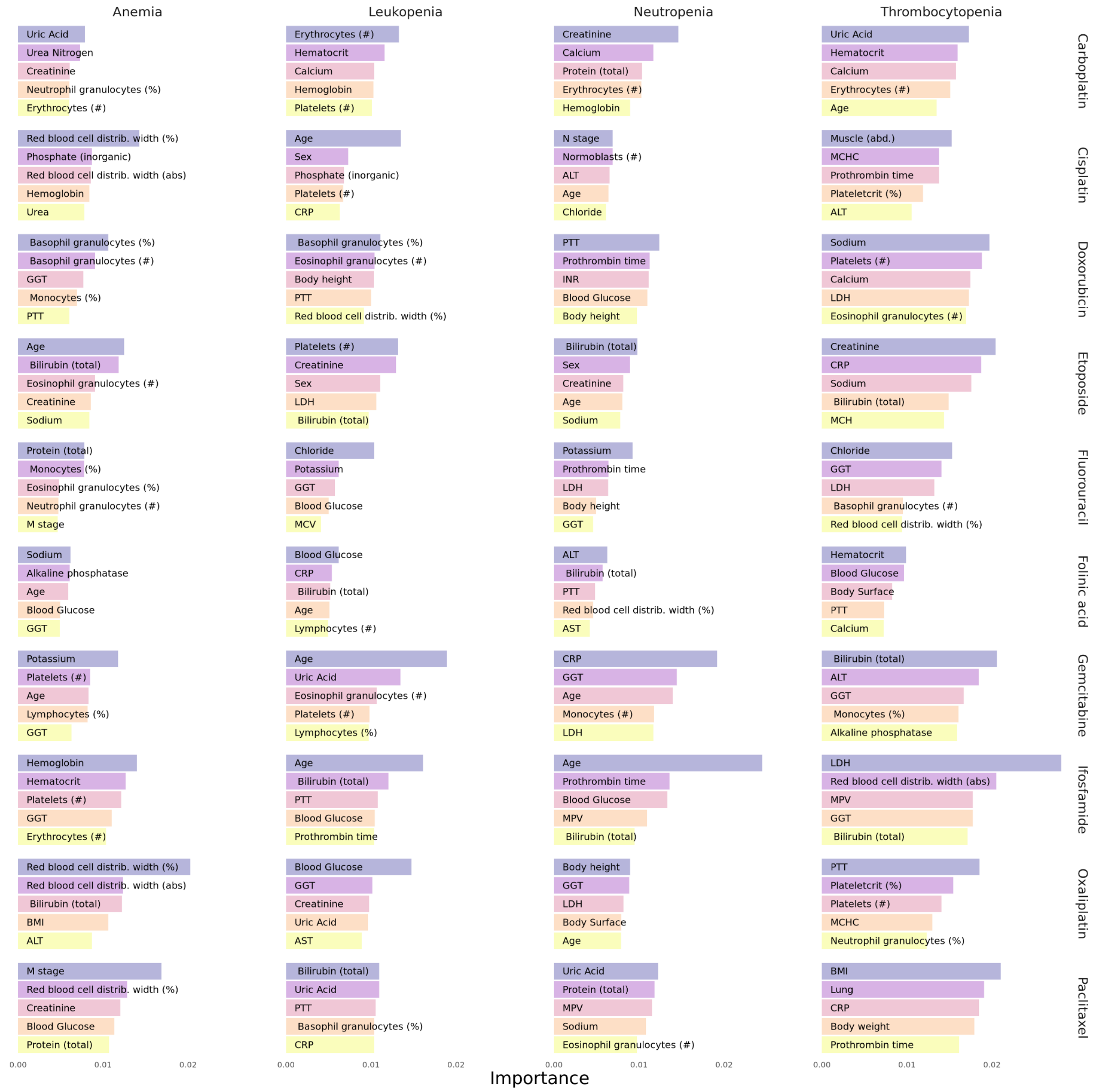


**Supplementary Figure 4:** Top 5 most important patient characteristics for adverse outcomes for the most frequently applied substances

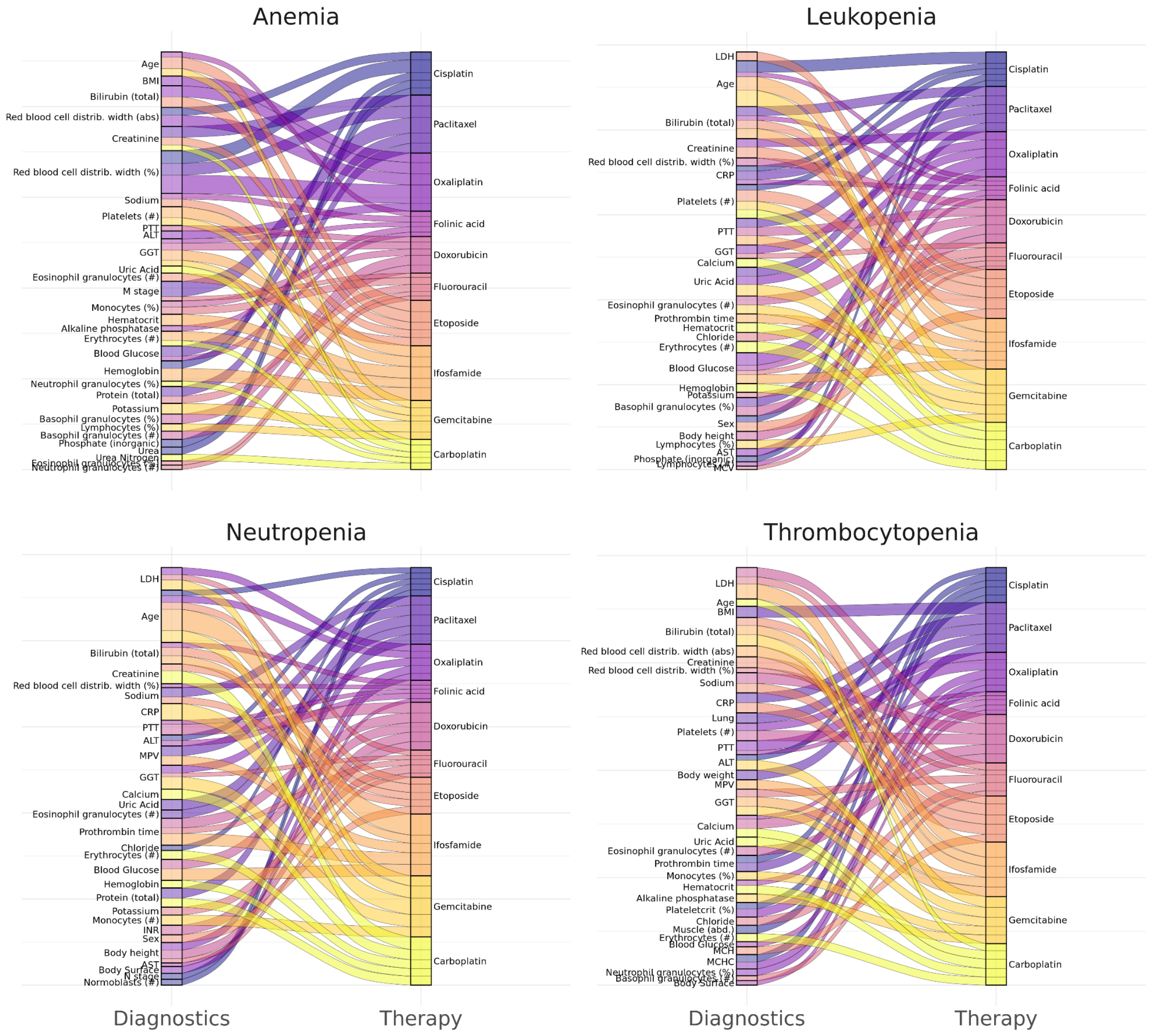


**Supplementary Figure 5**: Most important interactions between frequently applied cancer drugs and patient characteristics towards adverse hematological effects. The top 5 most important interactions for each substance are shown. Importance is defined as the mean of the absolute BiLRP scores.
